# Retinal Age as a Predictive Biomarker for Mortality Risk

**DOI:** 10.1101/2020.12.24.20248817

**Authors:** Zhuoting Zhu, Danli Shi, Guankai Peng, Zachary Tan, Xianwen Shang, Wenyi Hu, Huan Liao, Xueli Zhang, Yu Huang, Honghua Yu, Wei Meng, Wei Wang, Xiaohong Yang, Mingguang He

**Affiliations:** Department of Ophthalmology, Guangdong Academy of Medical Sciences, Guangdong Provincial People’s Hospital, Guangzhou, China; State Key Laboratory of Ophthalmology, Zhongshan Ophthalmic Center, Sun Yat-sen University, Guangzhou, China; Guangzhou Vision Tech Medical Technology Co., Ltd; Centre for Eye Research Australia; Ophthalmology, University of Melbourne, Melbourne, Australia; Neural Regeneration Group, Institute of Reconstructive Neurobiology, University of Bonn, Bonn, Germany; Ophthalmology, Department of Surgery, University of Melbourne, Melbourne, Australia

**Author notes:** **Corresponding author** Mingguang He, MD PhD, Xiaohong Yang, MD PhD, Wei Wang, MD PhD.

**Keywords:** retinal age, mortality, prediction

## Abstract

**Summary:** *Background:* Ageing varies substantially, thus an accurate quantification of ageing is important. We developed a deep learning (DL) model that predicted age from fundus images (retinal age). We investigated the association between retinal age gap (retinal age-chronological age) and mortality risk in a population-based sample of middle-aged and elderly adults.

*Methods:* The DL model was trained, validated and tested on 46,834, 15,612 and 8,212 fundus images respectively from participants of the UK Biobank study alive on 28^th^ February 2018. Retinal age gap was calculated for participants in the test (n=8,212) and death (n=1,117) datasets. Cox regression models were used to assess association between retinal age gap and mortality risk. A restricted cubic spline analyses was conducted to investigate possible non-linear association between retinal age gap and mortality risk.

*Findings:* The DL model achieved a strong correlation of 0·83 (P<0·001) between retinal age and chronological age, and an overall mean absolute error of 3·50 years. Cox regression models showed that each one-year increase in the retinal age gap was associated with a 2% increase in mortality risk (hazard ratio=1·02, 95% confidence interval:1·00-1·04, P=0·021). Restricted cubic spline analyses showed a non-linear relationship between retinal age gap and mortality (P_non-linear_=0·001). Higher retinal age gaps were associated with substantially increased risks of mortality, but only if the gap exceeded 3·71 years.

*Interpretation:* Our findings indicate that retinal age gap is a robust biomarker of ageing that is closely related to risk of mortality.

*Funding:* National Health and Medical Research Council Investigator Grant, Science and Technology Program of Guangzhou.

**Research in context:** *Evidence before this study:* Ageing at an individual level is heterogeneous. An accurate quantification of the biological ageing process is significant for risk stratification and delivery of tailored interventions. To date, cell-, molecular-, and imaging-based biomarkers have been developed, such as epigenetic clock, brain age and facial age. While the invasiveness of cellular and molecular ageing biomarkers, high cost and time-consuming nature of neuroimaging and facial ages, as well as ethical and privacy concerns of facial imaging, have limited their utilities. The retina is considered a window to the whole body, implying that the retina could provide clues for ageing.

*Added value of this study:* We developed a deep learning (DL) model that can detect footprints of aging in fundus images and predict age with high accuracy for the UK population between 40 and 69 years old. Further, we have been the first to demonstrate that each one-year increase in retinal age gap (retinal age-chronological age) was significantly associated with a 2% increase in mortality risk. Evidence of a non-linear association between retinal age gap and mortality risk was observed. Higher retinal age gaps were associated with substantially increased risks of mortality, but only if the retinal age gap exceeded 3·71 years.

*Implications of all the available evidence:* This is the first study to link the retinal age gap and mortality risk, implying that retinal age is a clinically significant biomarker of ageing. Our findings show the potential of retinal images as a screening tool for risk stratification and delivery of tailored interventions. Further, the capability to use fundus imaging in predicting ageing may improve the potential health benefits of eye disease screening, beyond the detection of sight-threatening eye diseases.

## Introduction

Globally, the population aged 60 and over is estimated to reach 2·1 billion in 2050.^1^ Ageing populations place tremendous pressure on health-care systems.^2^ The rate of ageing at an individual level is heterogeneous. An accurate quantification of the biological ageing process is significant for risk stratification and the delivery of tailored interventions.^3^

To date, several tissue-, cell-, molecular-, and imaging-based biomarkers have been developed, such as DNA-methylation status, brain age and three dimensional (3D) facial age.^4-7^ While the invasiveness of cellular and molecular ageing biomarkers, high cost and time-consuming nature of neuroimaging and facial ages, and ethical and privacy concerns of facial imaging, have limited their utilities.

The retina is considered a window to the whole body.^8-12^ In addition, the retina is amenable to rapid, non-invasive, and cost-effective assessments. The advent of deep learning (DL) has greatly improved the accuracy of image classification and processing. Recent studies have demonstrated successful applications of DL models in the prediction of age using clinical images.^5,6,13^ Taken together, this raises the potential that biological age can be predicted by applying DL to retinal images. For optimal utility, viable biomarkers of ageing must also relate to the risk of age-related morbidity and mortality.

We therefore developed a DL model that can predict age from fundus images, known as retinal age. Using a large population-based sample of middle-aged and elderly adults, we investigated the association between retinal age gap, defined as the difference between retinal age and chronological age, and all-cause mortality.

## Methods

### Study population

The UK Biobank is a large-scale, population-based cohort of more than 500,000 UK residents aged 40-69 years. Participants were recruited between 2006 and 2010, with all participants completing comprehensive health-care questionnaires, detailed physical measurements, and biological sample collections. Health-related events were ascertained via data linkage to hospital admission records and mortality registry. Ophthalmic examinations were introduced in 2009. The overall study protocol and protocols for each test have been described in extensive details elsewhere.^14^

The National Information Governance Board for Health and Social Care and the NHS North West Multicenter Research Ethics Committee approved the UK Biobank study (11/NW/0382) in accordance with the principles of the Declaration of Helsinki, with all participants providing informed consent. The present analysis operates under UK Biobank application 62525.

### Fundus photography

Ophthalmic measurements including LogMAR visual acuity, autorefraction and keratometry (Tomey RC5000, Tomey GmbH, Nuremberg, Germany), intraocular pressure (IOP, Ocular Response Analyzer, Reichert, New York, USA), and paired retinal fundus and optical coherence tomography imaging (OCT, Topcon 3D OCT 1000 Mk2, Topcon Corp, Tokyo, Japan) were collected. A 45-degree non-mydriatic and non-stereo fundus image centered to include both the optic disc and macula was taken for each eye. A total of 131,238 images from 66,500 participants were obtained from the UK Biobank study, among which 80,170 images from 46,970 participants passed the image quality check.

### Deep learning model for age prediction

To build the DL model for age prediction, participants from the UK Biobank study alive on 28^th^ February 2018 (N_subj_=46,970) were randomly split into three datasets – training (N_subj_=27,424, 60% of participants), validation (N_subj_=9,142, 20%), and test (N_subj_=9,142, 20%). For the training and validation datasets, fundus images from both eyes (if available) were used to maximise the volume of data available (N_img_=46,834 and 15,612 respectively). For the test dataset, fundus images from right eyes were selected for primary analyses (N_subj_=8,212), while fundus images from left eyes were selected for sensitivity analyses.

The development and validation of the DL model for age prediction are outlined in Figure 1. Briefly, all fundus images were preprocessed by subtracting average color,^15^ resized to a resolution of 299*299 pixels, and pixel values rescaled to 0∼1. After preprocessing, images were fed into a DL model using a Xception architecture. During training, data augmentation was performed using random horizontal or vertical flips and the algorithm optimised using stochastic gradient descent. To prevent overfitting, we implemented a dropout of 0·5, and carried out early stopping when validation performance did not improve for 10 epochs. The selection of DL models was based on performance in the validation set. The performance of the DL model, including mean absolute error (MAE) and correlation between predicted retinal age and chronological age, was calculated. We then retrieved attention maps from the DL models using guided Grad-CAM,^16^ which highlights pixels in the input image based on their contributions to the final evaluation.

**Figure 1.**
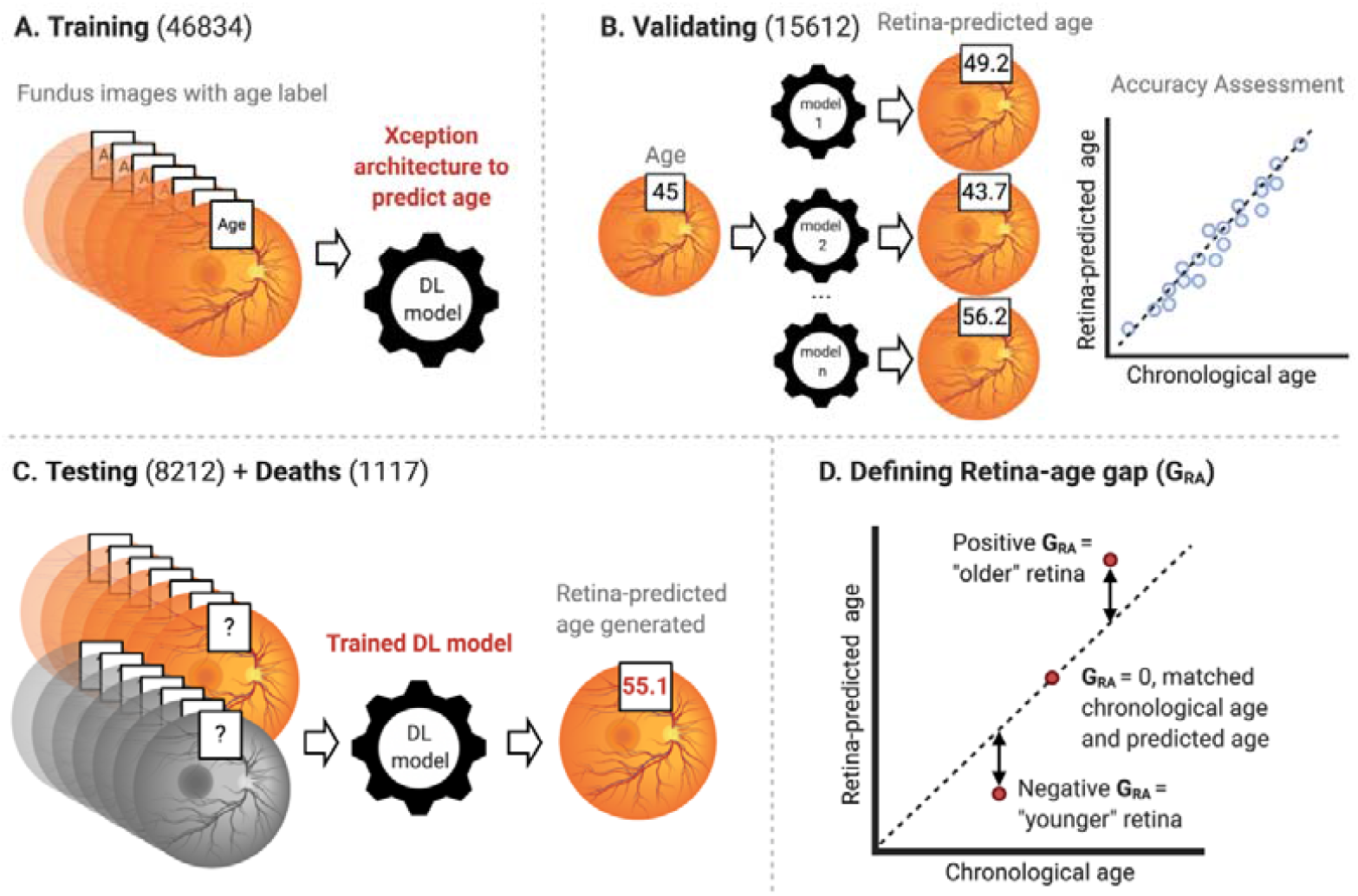
Overview of the study workflow. Figures showing the study workflow used to calculate retinal age gaps from fundus images. Fundus images were preprocessed and fed into the DL model. (A) The Xception architecture was used to train fundus images, with chronological age as the outcome variable; (B) The selection of DL models was based on performance in the validation set, where predicted retinal and chronological ages were compared; (C) The selected trained DL model was then applied to make retinal age predictions from fundus images for participants in the test and death datasets; (D) The difference between predicted retinal age and chronological age was defined as the retinal age gap. A positive retinal age gap indicated an ‘older’ appearing retina, while a negative retinal age gap indicated a ‘younger’ appearing retina. This figure was created with BioRender.com.

### Retinal age gap definition

The difference between retinal age predicted by the DL model and chronological age was defined as the retinal age gap. A positive retinal age gap indicated an ‘older’ appearing retina, while a negative retinal age gap indicated a ‘younger’ appearing retina.

### Mortality ascertainment

Mortality status and date of death were ascertained via data linkage to the National Health Service central mortality registry. Participants who had died from all causes during the follow-up period (N_subj_=1,117) were included in the death dataset. Duration of follow-up for each participant (person-year) was calculated as the length of time between baseline age and date of death, loss to follow-up, or complete follow-up (28^th^ February 2018), whichever came first.

### Covariates

Factors previously known to be associated with mortality^17^ were included as potential confounders in the present analyses. These variables included baseline age, sex, ethnicity (recorded as white and non-white), Townsend deprivation indices (an area-based proxy measure for socioeconomic status), education attainment (recorded as college or university degree, and others), smoking status (recorded as current/previous and never), physical activity level (recorded as above moderate/vigorous/walking recommendation and not), general health status (recorded as excellent/good and fair/poor), and comorbidities (obesity, diabetes mellitus, hypertension, history of heart diseases, and history of stroke).

Body mass index (BMI) was calculated as body weight in kilograms divided by height squared. Obesity was defined as BMI ≥30 kg/m^2^. Diabetes mellitus was defined as self-reported or doctor-diagnosed diabetes mellitus, the use of anti-hyperglycaemic medications or insulin, or a glycosylated haemoglobin level of ≥6·5%. Hypertension was defined as self-reported, or doctor-diagnosed hypertension, the use of antihypertensive drugs, an average systolic blood pressure of at least 130mmHg or an average diastolic blood pressure of at least 80mmHg. Self-reported history of angina and heart attack was used to classify history of heart diseases.

### Statistical analyses

Descriptive statistics, including means and standard deviations (SDs), numbers and percentages, were used to report baseline characteristics of study participants. The retinal age gap was calculated for participants in the test and death datasets, and further used to explore the association between retinal age gap and mortality risk. Cox proportional hazards regression models considering retinal age gap as a continuous linear term were fitted to estimate the effect of a one-year increase in retinal age gap on mortality risk. We then investigated associations between retinal age gaps at different quantiles with mortality. In addition, a restricted cubic spline analyses of possible non-linear associations between retinal age gap and mortality status was performed, with 5 knots placed at equal percentiles of the retinal age gap, and retinal age gap of zero years used as the reference value. We adjusted Cox models for the following covariates – baseline age, sex, ethnicity, and Townsend deprivation indices (model I); additional educational level, obesity, smoking status, physical activity level, diabetes mellitus, hypertension, general health status, history of heart diseases, and history of stroke (model II).

The proportional hazards assumption for each variable included in the Cox proportional hazards regression models were graphically assessed. All variables were found to meet the assumption. A two-sided p value of < 0·05 indicated statistical significance. Analyses were performed using R (version 3.3.0, R Foundation for Statistical Computing, www.R-project.org, Vienna, Austria) and Stata (version 13, StataCorp, Texas, USA).

## Results

### Study sample

The study population characteristics are described in Table 1. The DL model was trained and validated on subsets of participants with mean ages of 55·6 ± 8·21 and 55·7 ± 8·19 years; and with 55·9% and 55·2% female, respectively. For the test and death datasets, participants had mean ages of 55·5 ± 8·22 and 61·0 ± 6·67 years; and were 55·1% and 42·2% female, respectively.

**Table 1.** Characteristics of datasets derived from the UK Biobank study

|  | Training <sup>a</sup> | Validation <sup>a</sup> | Test | Death |
| --- | --- | --- | --- | --- |
| N<sub>subj</sub> | 27424 | 9142 | 8212 | 1117 |
| N<sub>img</sub> | 46834 | 15612 | 8212 | 1117 |
| Mean age (mean±SD, yrs) <sup>b</sup> | 55·6±8·21 | 55·7±8·19 | 55·5±8·22 | 61·0±6·67 |
| Female, N (%) <sup>b</sup> | 15,341 (55·9) | 5,045 (55·2) | 4,526 (55·1) | 471 (42·2) |
| White ethnicity, N (%) <sup>b</sup> | 25,400 (92·6) | 8,468 (92·6) | 7,618 (92·8) | 1,035 (92·7) |
| Townsend index (mean±SD) <sup>b</sup> | -1·09±2·94 | -1·10±2·96 | -1·08±2·95 | -0·73±3·12 |
| College or university degree, N (%) <sup>b</sup> | 9,981 (36·4) | 3,344 (36·6) | 3,035 (37·0) | 299 (26·8) |
| Current/previous smoker, N (%) <sup>b</sup> | 11,671 (42·8) | 3,905 (43·0) | 3,532 (43·1) | 659 (59·4) |
| Above physical activity recommendation, N (%) <sup>b</sup> | 18,746 (82·7) | 6,219 (82·4) | 5,682 (83·4) | 681 (77·4) |
| Excellent/good health status, N (%) <sup>b</sup> | 20,290 (74·5) | 6,738 (74·0) | 6,149 (75·2) | 618 (55·8) |
| Obesity, N (%) <sup>b</sup> | 6,239 (22·9) | 2,112 (23·2) | 1,840 (22·5) | 312 (28·1) |
| Diabetes mellitus, N (%) <sup>b</sup> | 1,332 (4·86) | 443 (4·85) | 428 (5·21) | 133 (11·9) |
| Hypertension, N (%) <sup>b</sup> | 19,632 (71·6) | 6633 (72·6) | 5,956 (72·5) | 928 (83·1) |
| History of heart diseases, N (%) <sup>b</sup> | 848 (3·09) | 296 (3·24) | 250 (3·04) | 94 (8·42) |
| History of stroke, N (%) <sup>b</sup> | 309 (1·13) | 83 (0·91) | 103 (1·25) | 35 (3·13) |

N<sub>subj</sub> = number of subjects; N<sub>img</sub> = number of images; yrs = years; SD = standard deviation.
<sup>a</sup> Selection of images of both eyes if available.
<sup>b</sup> Values are based on N<sub>subj</sub>.

### Deep learning model performance for age prediction

Figure 2A shows the performance of the DL model on the test dataset. The trained DL model was able to achieve a strong correlation of 0·83 (P<0·001) between predicted retinal age and chronological age, with an overall MAE of 3·50 years. Two representative examples of fundus images with corresponding attention maps for age prediction are shown in Figure 3. Regions around retinal vessels are highlighted by the DL model for age prediction.

**Figure 2.**
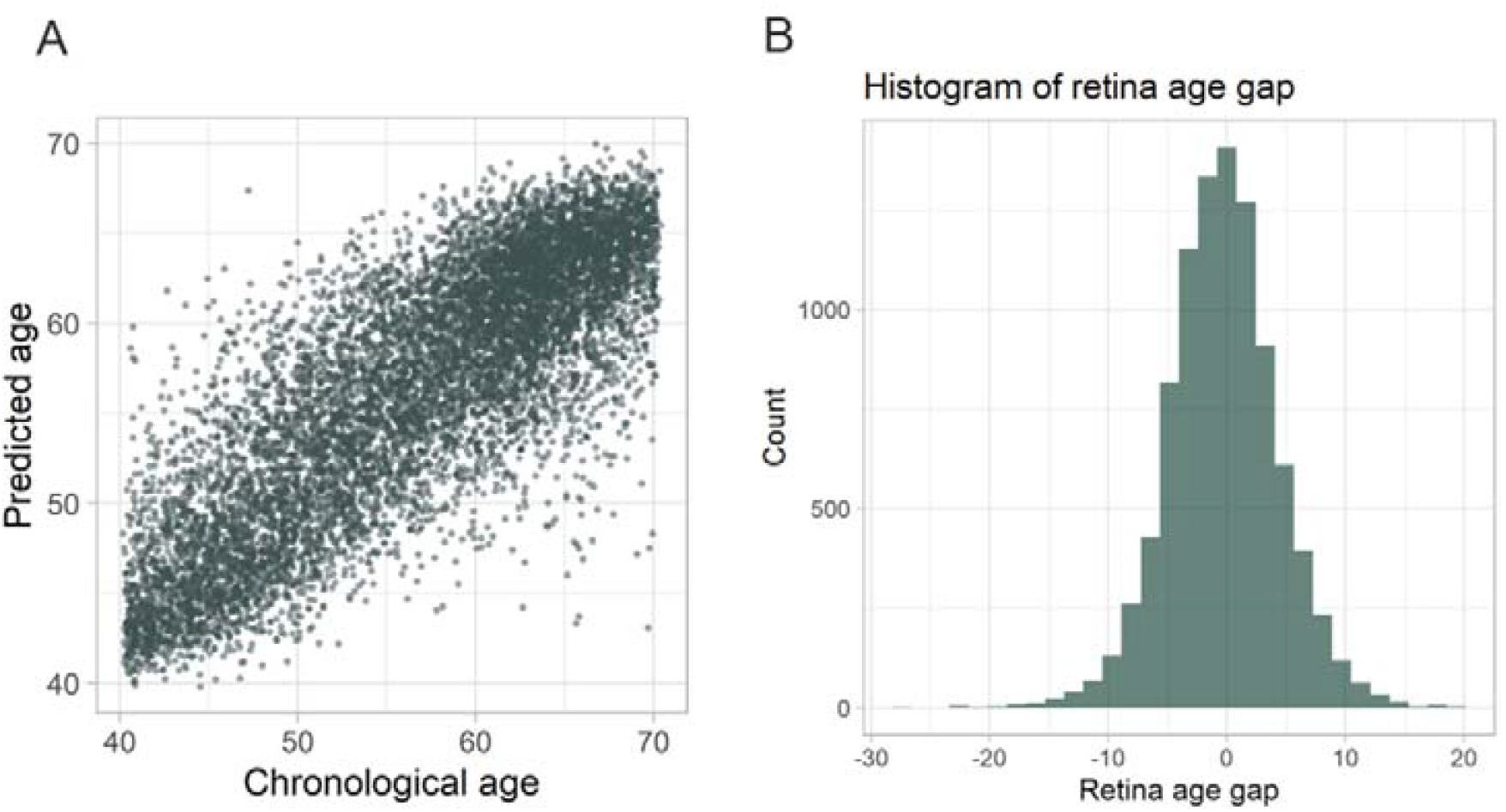
Performance of the deep learning model on the test dataset. (A) Scatterplot depicting correlation of predicted age (y-axis) with chronological age (x-axis); (B) Histogram showing the nearly normal distribution of the retinal age gap.

**Figure 3.**
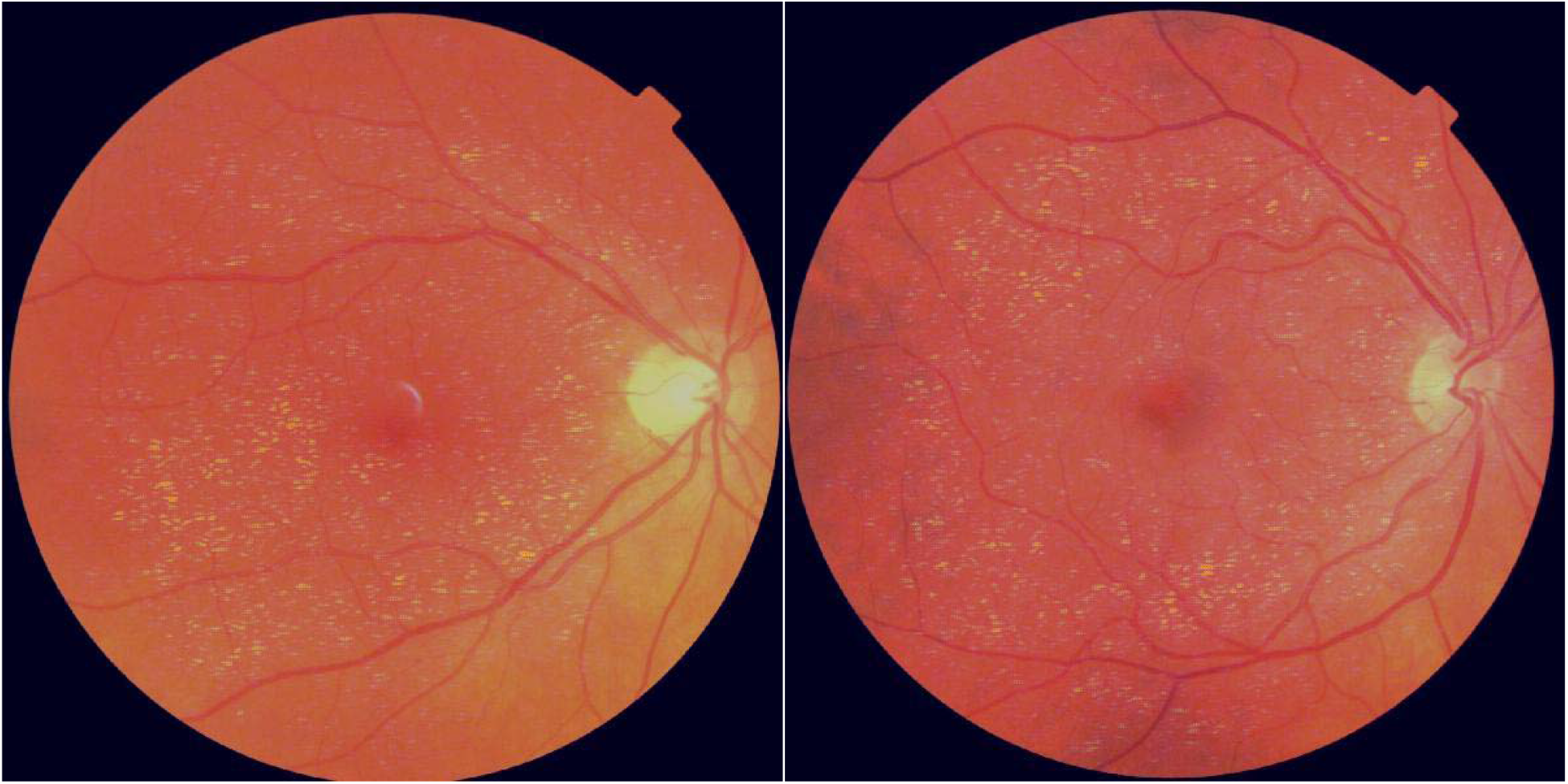
Attention maps for age prediction. Figures showing representative examples of fundus images with corresponding attention maps for age prediction. Regions highlighted with a brighter colour indicate areas that are used by the DL model for age prediction. Regions around the retinal vessels are highlighted.

### Retinal age gap

The distribution of the retinal age gap followed a nearly normal distribution (Figure 2B). The mean (SD) and median (interquartile range) of the retinal age gap were −0.16 (4.54) and −0.19 (−2.99, 2.60). The proportions of fast agers with retinal age gaps more than 3, 5 and 10 years were 22.0%, 12.0% and 1.67%, respectively.

### Retinal age gap and mortality

Considering linear effects only and following adjustment for all confounding factors, each one-year increase in retinal age gap was associated with a 2% increase in mortality risk (hazard ratio [HR] = 1·02, 95% confidence interval [CI]: 1·00-1·04, P = 0·021; Table 2). Compared to participants with retinal age gaps in the lowest quantile, mortality risk was comparable for those in the second and the third quantiles (HR = 1·05, 95% CI: 0·88-1·24, P = 0·602; HR = 0·89, 95% CI: 0·73-1·09, P = 0·261, respectively). Mortality risk was significantly increased for participants with retinal age gaps in the fourth quantile (HR = 1·33, 95% CI: 1·06-1·67, P = 0·012; Table 2).

**Table 2.**
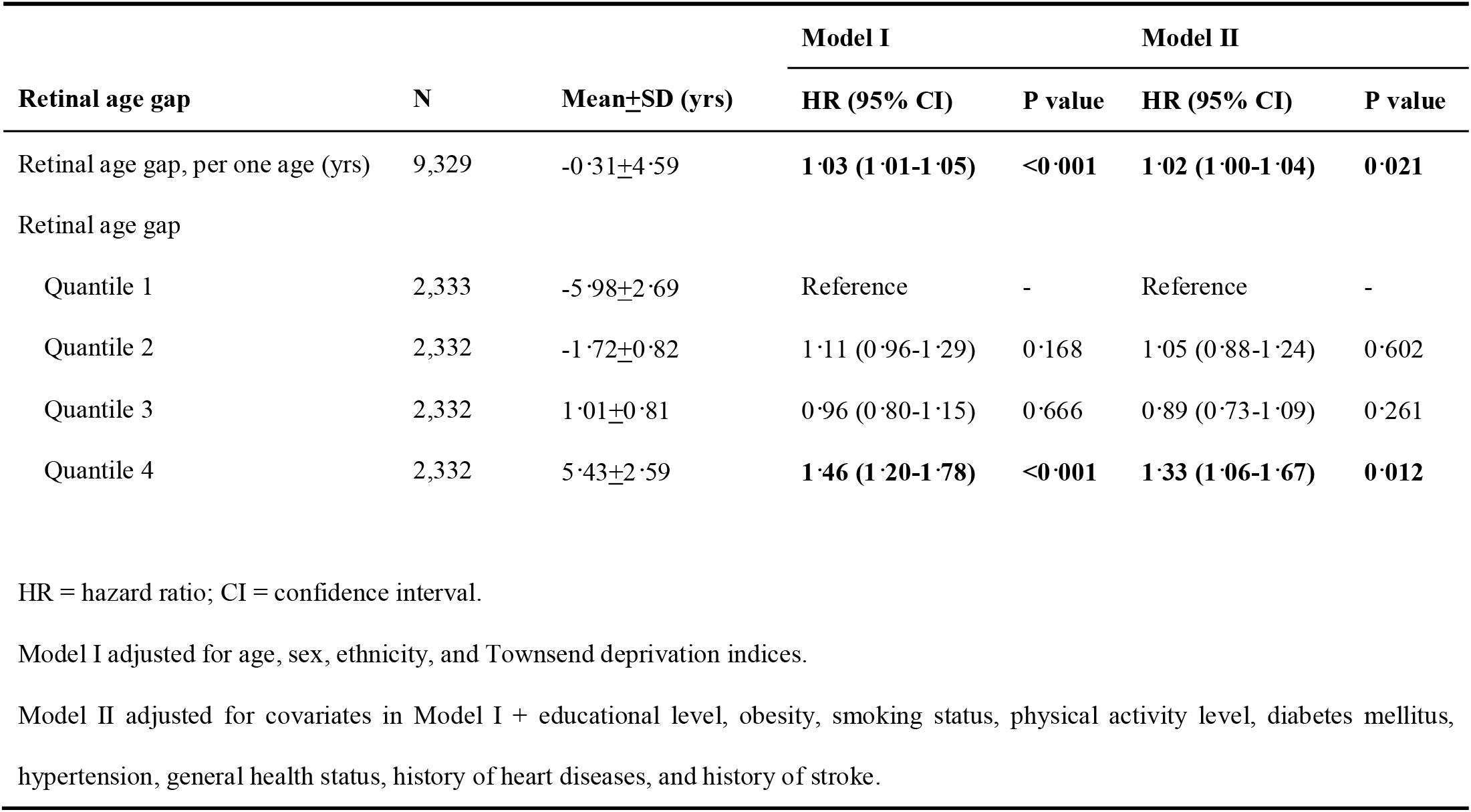
Association between retinal age gap with mortality using Cox proportional hazards regression models

Allowing for non-linearity, Figure 5 illustrates the estimated association between retinal age gap and mortality risk. Evidence of an overall and non-linear association between retinal age gap and mortality risk was observed (P_overall_ < 0·001; P_non-linear_ = 0·001). Higher retinal age gaps were associated with substantially increased risks of mortality, but only if the retinal age gap exceeded 3·71 years.

**Figure 4.**
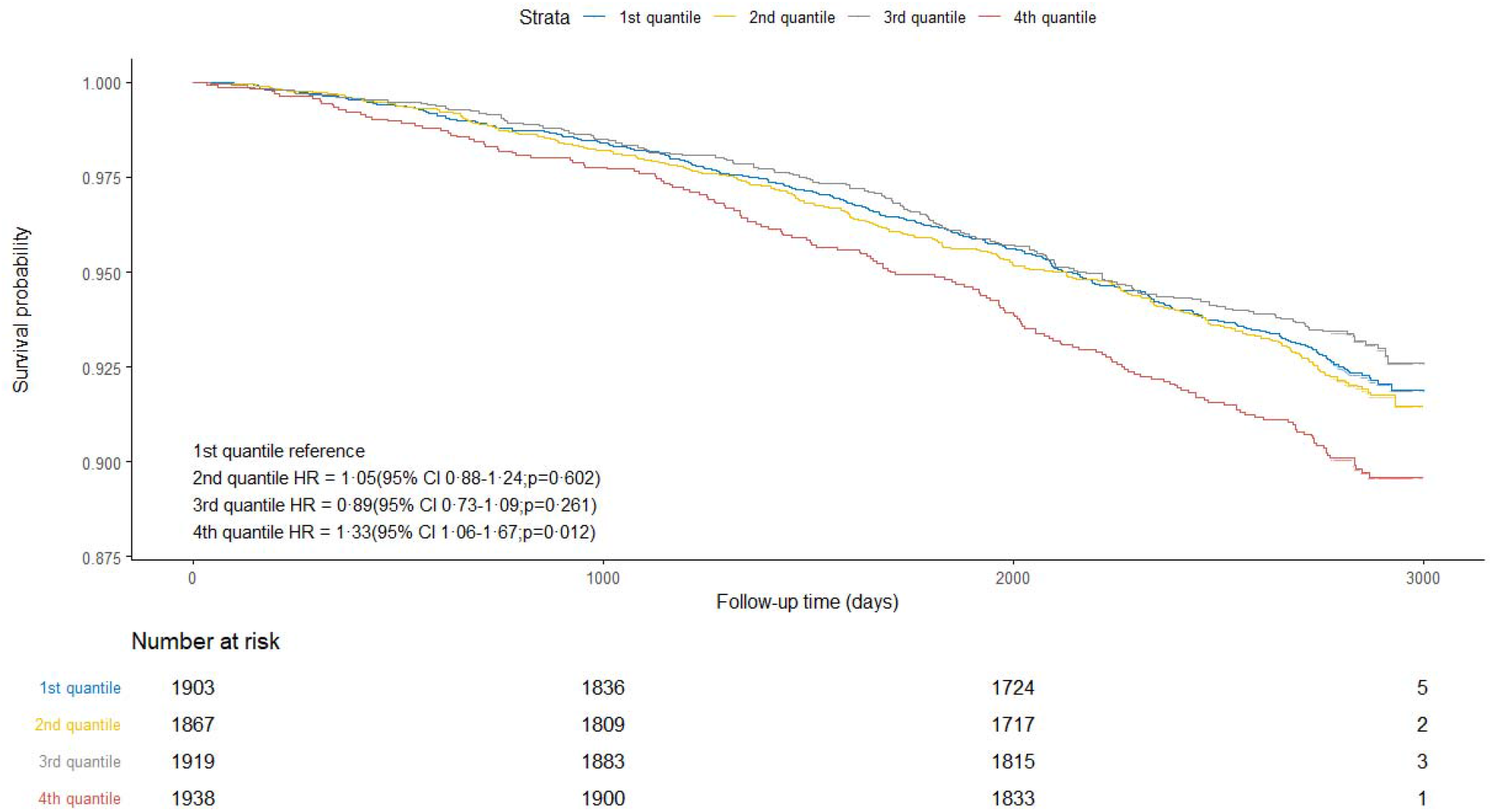
Adjusted survival curves for mortality risk by retinal age gap quantiles. Mortality risk is shown over time for participants in different retinal age gap quantiles. Lower quantiles corresponded to participants who had chronological ages greater than predicted retinal age, whereas higher quantiles corresponded to those with chronological ages lower than predicted retinal age. Plots were based on Cox proportional hazards regression models, adjusted for age, sex, ethnicity, Townsend deprivation indices, educational level, obesity, smoking status, physical activity level, diabetes mellitus, hypertension, general health status, history of heart diseases, and history of stroke. Compared to participants with retinal age gaps in the lowest quantile, mortality risk was comparable for those in the second and the third quantiles (hazard ratio [HR] = 1·05, 95% confidence interval [CI]: 0·88-1·24, P = 0·602; HR = 0·89, 95% CI: 0·73-1·09, P = 0·261, respectively). Mortality risk was significantly increased for participants with retinal age gaps in the fourth quantile (HR = 1·33, 95% CI: 1·06-1·67, P=0·012).

**Figure 5.**
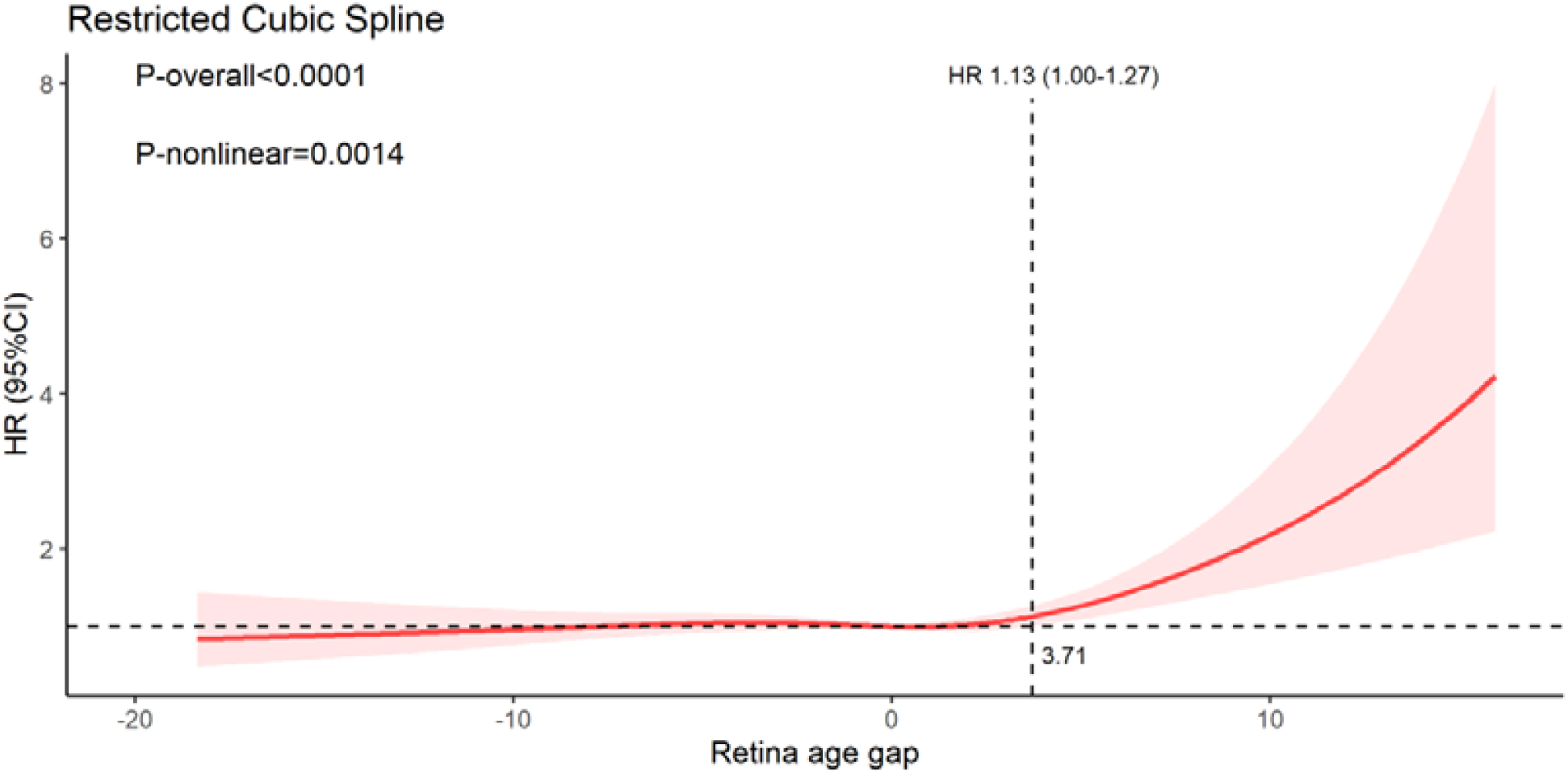
Association between retina age gap and mortality risk, allowing for non-linear effects. The reference retinal age gap for this plot (with hazard ratio [HR] fixed as 1·0) was 0 years. The model was fitted with a restricted cubic spline for retinal age gap (knots placed at equal percentiles of retina age gap), adjusted for age, sex, ethnicity, Townsend deprivation indices, educational level, obesity, smoking status, physical activity level, diabetes mellitus, hypertension, general health status, history of heart diseases, and history of stroke. Evidence of an overall and non-linear association between retinal age gap and all-cause mortality was observed (P_overall_ < 0·001; P_non-linear_ = 0·001). Higher retinal age gaps were associated with substantially increased risks of mortality, but only if the retinal age gap exceeded 3·71 years.

### Sensitivity analyses

In order to verify the robustness of our findings, fundus images from left eyes were chosen for the statistical analyses. Similar results were observed for left eyes (data not shown).

## Discussion

Using a large population-based sample of middle-aged and elderly adults, we developed a DL model that could predict age from fundus images with high accuracy. Further, we found that the retinal age gap, defined as the difference between predicted retinal age and chronological age, independently predicted the risk of mortality. Our findings have demonstrated that retinal age is a robust biomarker of ageing that can predict all-cause mortality.

To the best of our knowledge, this is the first study that has proposed retinal age as a biomarker of ageing. Our trained DL model achieved excellent performance with a MAE of 3·5, outperforming most existing biomarkers in the prediction of age. Previous studies have demonstrated MAEs of 3·3-5·2 years for DNA methylation clock,^18,19^ 5·5-5·9 years MAEs for blood profiles,^20,21^ and 6·2-7·8 years MAEs for the transcriptome ageing clock.^22,23^ Neuroimaging and 3D facial imaging have achieved accurate performances in age prediction with MAEs between 4·3 and 7·3,^7,24^ and 2·8 and 6·4 years,^6,25^ respectively. Despite these reasonable accuracies, the invasiveness of cellular and molecular ageing biomarkers, high cost and time-consuming nature of neuroimaging and 3D facial ages, and ethical and privacy concerns of facial imaging, have limited their utilities. In addition to excellent performance in age prediction, determining retinal age using fundus images is fast, safe, cost-effective and user-friendly, thus offering great potential for use in a large number of people.

Beyond age prediction, our study has extended the application of retinal age to the prediction of survival. Our novel findings have determined that the retinal age gap is an independent predictor of increased mortality risk, further suggesting that retinal age is a clinically significant biomarker of ageing. The relevance of retinal age for general health is intuitive, given that the retina is the only organ that is amenable to *in vivo* visualisation of the microvasculature and neural tissue. The retina offers a unique, accessible ‘window’ to evaluate underlying pathological processes of systemic vascular and neurological diseases that are associated with increased risks of mortality. This hypothesis is supported by previous studies which have suggested that retinal imaging contains information about cardiovascular risk factors,^26^ chronic kidney diseases^27^ and systemic biomarkers.^28^ In addition, this hypothesis is also consistent with previously reported qualitative and quantitative studies that have found that ocular imaging measures (e.g. retinal-vessel calibre) and retinal diseases (e.g. glaucoma) are significantly associated with mortality.^29,30^ This body of work supports the hypothesis that the retina plays an important role in the ageing process and is sensitive to the cumulative damages of ageing which increase the mortality risk.

Our findings have several important clinical implications. Firstly, the fast, non-invasive, and cost-effective nature of fundus imaging enables it to be an accessible screening tool to identify individuals at an increased risk of mortality. This risk stratification will assist tailored health-care decision-making, as well as targeting and monitoring of interventions. Given the rising burden of non-communicable diseases and population ageing globally, the early identification and delivery of personalised health-care may have tremendous public health benefits. Further, the recent development of smartphone-based retinal cameras, together with the integration of DL algorithms, may in the future provide point-of-care assessments of ageing and improve accessibility to tailored risk assessments. Secondly, the capability to use fundus images in predicting ageing may improve potential health benefits of eye disease screening, beyond the diagnosis of sight-threatening eye diseases. This may improve the health economic cost-effectiveness of programs such as diabetic retinopathy screening, thus increasing the impact and access to eye disease screening programs.

The large-scale sample size, long-term follow-up, standardised protocol in capturing fundus images, validity of mortality data, and adjustment for a wide range of confounding factors in the statistical models of this study support the robustness of our findings. Despite these promising results, our study has several limitations. Firstly, these current analyses are limited by retinal images that were captured at a particular cross-section in time, with trajectories in retinal ageing potentially being a better indicator of mortality. Secondly, participants involved in the UK Biobank study were volunteers, who might not be representative of the population from which they were drawn. Of note, the potential healthy effect might underestimate effects of retinal age gap on mortality, as individuals with extremely poor health were less likely to participate in this study. Thirdly, the lack of external datasets might limit the generalisability of our DL algorithms and findings. Lastly, we were unable to fully exclude the possibility of residual confounders between retinal age gap and mortality.

## Conclusion

In summary, we have developed a DL algorithm that can detect footprints of ageing in fundus images and predict age with high accuracy. Further, we have been the first to demonstrate that the retinal age gap is significantly associated with an increased risk of mortality. Our findings suggest that retinal age is a robust biomarker of ageing. Lastly, our work calls for future research into applications of the retinal age gap, and whether retinal age can be used to better understand processes underpinning ageing.

## Data Availability

The authors confirm that the data supporting the findings of this study are available within the article.

## Contributors

ZZ and SD conceptualised and designed the study with WW, HM, and YX. ZZ and SD did the literature search and wrote the first draft of the manuscript. SD, PG and MW did the deep learning modelling, ZZ, SX and WW did the statistical analysis.

WW, HM and YX had full access to all of the data. All authors commented on the manuscript.

## Acknowledgments

This present work was supported by the NHMRC Investigator Grant (APP1175405), Fundamental Research Funds of the State Key Laboratory of Ophthalmology, National Natural Science Foundation of China (82000901), Project of Investigation on Health Status of Employees in Financial Industry in Guangzhou, China (Z012014075), Science and Technology Program of Guangzhou, China (202002020049). Professor Mingguang He receives support from the University of Melbourne through its Research Accelerator Program and the CERA Foundation. The Centre for Eye Research Australia (CERA) receives Operational Infrastructure Support from the Victorian State Government.

